# Repeat Testing of Mpox Specimens with Late CTs Improves Detection of Potential False Positive Cases

**DOI:** 10.1101/2023.10.03.23296486

**Authors:** Ryan C. Shean, Weston C. Hymas, Jeremy Klein, Michael Pyne, David R. Hillyard, Benjamin T. Bradley

## Abstract

The global outbreak of mpox necessitated the rapid development of clinical assays for monkeypox virus detection. While the majority of mpox specimens have high viral loads with corresponding early CT values, reports have indicated some specimens with late CT values can represent false positive results. To mitigate this risk, the Centers for Disease Control and Prevention (CDC) published an advisory recommending repeat testing of all specimens with CT values ≥34. However, limited experimental data was available to support this specific cutoff. In this study, we examine whether a more conservative approach in which all specimens with CT values ≥29 are repeated would improve the detection of potential false positive results. Compared to the CDC algorithm, our approach identified an additional 20% (5/25) of potential false positive results. To assess the impact of this cutoff on laboratory workload, we estimated the expected increase in test volume and turnaround time (TAT) relative to the CDC method. Using a lower repeat threshold, test volume increased by 0.7% and the mean TAT for positive specimens increased by less than 15 minutes. Overall, a lower threshold than recommended by the CDC for repeating late CT mpox specimens may reduce the number of false positives reported while minimally impacting testing volume and TAT.

## Introduction

Monkeypox virus (MPXV) is a double stranded DNA virus in the *Orthopoxvirus* genus, which also includes variola virus, the causative agent of smallpox. In July 2022 the World Health Organization declared a Public Health Emergency of International Concern due to an outbreak of MPXV with sustained human-to-human transmission in multiple non-endemic countries^1^. At the beginning of the outbreak, testing capacity in the United States was limited to the Centers for Disease Control and Prevention (CDC) and members of the Laboratory Response Network. In June 2022, clinical laboratories, using the assay developed by the CDC, began adding additional testing capacity^2^. This was followed over the subsequent weeks by other clinical microbiology laboratories that either adapted the CDC assay or validated their own laboratory developed test in accordance with recommendations from the FDA^3^. Additionally, several manufacturers received emergency use authorization (EUA) for orthopoxvirus or MPXV-specific assays following the EUA declaration on September 17^th^, 2022.

The majority of specimens collected from individuals with mpox have demonstrated high viral loads with corresponding early CT values^4^. However, shortly after the outbreak began, reports of false positive mpox cases began to surface. In a series of three false positive cases, all specimens had late CT values ranging from 34.3 to 36.7^5^. In September 2022, the CDC published an advisory recommending repeat testing of all specimens with CT values ≥34^6^. Due to the limited data surrounding the utility of this cutoff, our institution adopted a more conservative strategy of repeating all positive specimens with a CT ≥29 from 9/15/22 to 11/4/22. Here we present a retrospective analysis of this data to better understand the clinical and laboratory impact of a more stringent repeat threshold.

## Materials and Methods

### MPXV PCR testing

Detection of monkeypox virus at ARUP was performed as previously described ^4^. In brief, specimens received as dry swabs were resuspended in PBS prior to further processing. DNA from VTM and dry swabs was extracted and amplified on the Roche cobas 6800 platform. Roche omni channel reagents with lab-developed primers and probes were used for detection of a conserved region of the polymerase gene present in non-variola orthopoxviruses^7^.

### Data extraction and labeling

Data for individual specimens were extracted from the Laboratory Information System (LIS) at the end of the study period. All tests with a completed time logged in the LIS between 00:00 on 9/15/22 and 23:59 on 11/4/22 were included. Information gathered for each specimen included a unique deidentified specimen ID, unique deidentified patient ID, age and sex of patient, initial CT value (CT1), CT value of repeated test if performed (CT2), swab type (Dry, VTM, or Other), collection site, collection time, in-lab time, and completion time. Samples were retrospectively classified based on their CT1 value as “detected” or “not detected”. “Detected” specimens were samples with CT1 <50. “Not detected” specimens were samples with CT1 = 50 (value provided by the instrument when no amplification curve is detected). “Detected” specimens were further sub-categorized as “positive”, “low positive”, or “indeterminate” based on CT1 and CT2 results. “Positive” specimens were samples with CT1 <29. “Low positive” specimens were samples with CT1 values between ≥29 and <50 and CT2 values <50. “Indeterminate” specimens also had CT1 values between ≥29 and <50 but with a CT2 value = 50. Nine specimens had their CT2 values excluded because the CT1 values were <29. These samples were repeated due to run control failures or when collected from pediatric patients (all pediatric specimens were repeated regardless of CT value). The impact of repeat testing on pediatric specimens was previously examined^8^. 21 specimens with a CT1 ≥29 failed to undergo repeat testing due to low specimen volume. These were classified as “detected” and excluded from sub-categorization.

### Ethics, data analysis, and statistics

University of Utah granted an IRB exemption ID#00158025. Deidentified data was processed in Python 3.8 and detailed statistical analysis was performed in R v4.2^9^.

## Results

### Descriptive epidemiology of MPXV PCR testing

A total of 4,233 specimens were analyzed over the 51-day study period with an average of 88.2 tests per day (Fig. 1). During this period, MPXV was initially detected in 18.7% (790/4,233) of specimens. Of the 790 specimens in which MPXV was detected 23.3% (184/790) had a CT1 at or above the repeat threshold of CT 29 and were repeated. Of the 184 tests that were repeated 86.4% (159/184) produced a valid CT2 signal and were classified as “low positive”. 13.6% (25/184) of these specimens failed to repeat and were classified as “indeterminate”. Total testing volume decreased by an average of 1.8 tests/day over the study period, however, detection rate and rate of low positives remained relatively stable.

**Figure 1.**
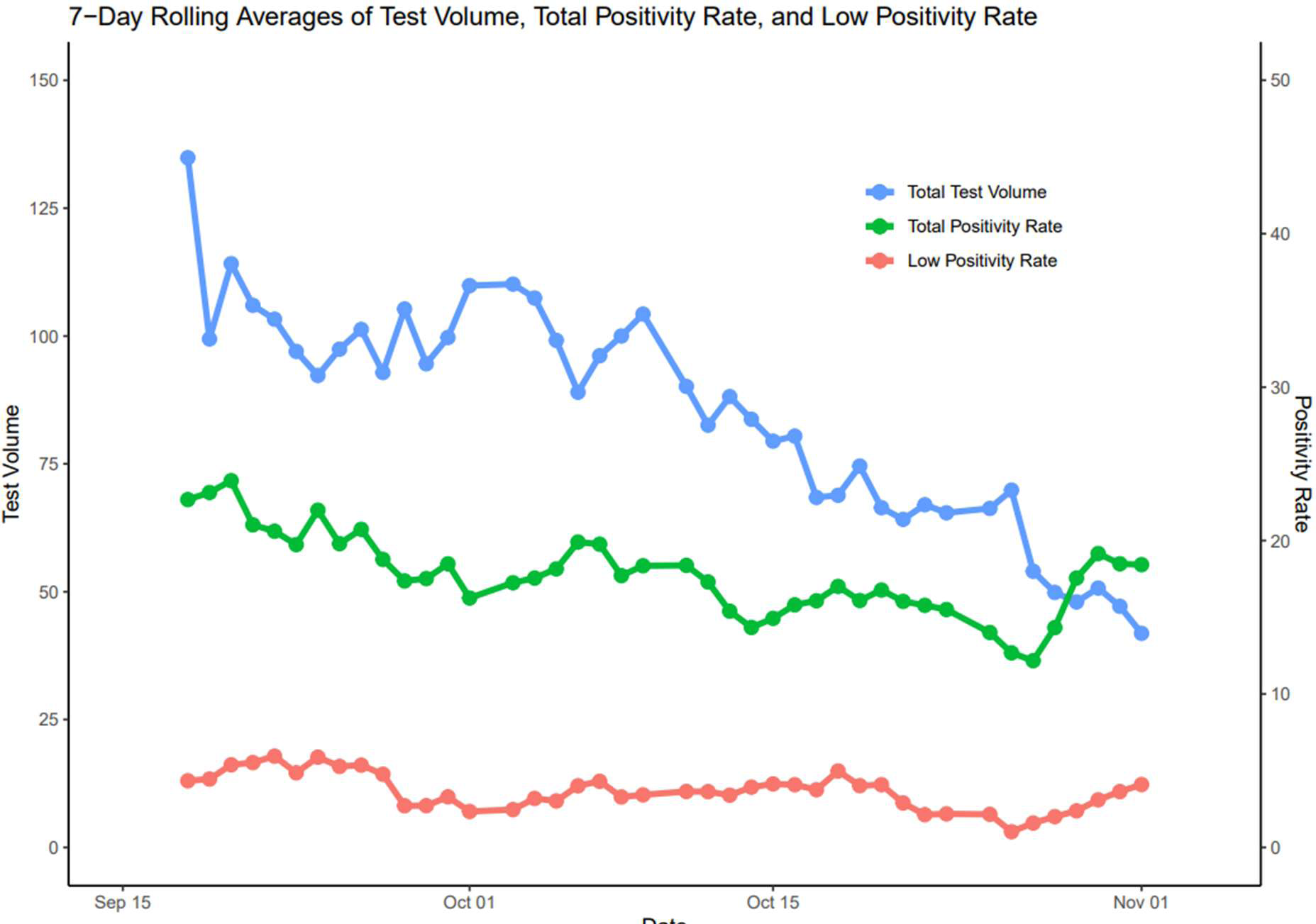
7-Day Rolling Averages of Test Volume, Total Positivity Rate, and Low Positivity Rate Total test volume is represented by the blue line and measured in tests per day on the left Y-axis. Percent of total tests with MPXV detected (CT1 <50) and percent of specimens falling within the repeat threshold (CT1 ≥29 and <50) are indicated by the green and red lines, respectively. Positivity rates are measured relative to the total test volume and mapped to the right Y-axis.

Specimens were collected from 3,111 individuals over the study period (Table 1). The average age of tested individuals was 34.4 years (SD 17.0) and 68.2% of patients were male. Overall, there was a significant association between categorical result and sex (Pearson’s Chi-squared test p<0.05). In cross-group comparisons, male individuals comprised a smaller proportion of negative results (62.7%) than either positive (93.0%) or low positive (92.8%) results (pairwise Bonferroni corrected Pearson’s chi-squared tests p<0.008). There was no significant association between age of patient and result.

**Table 1.** Patient characteristics by MPXV detection result.

| Result | Patients (n, %) | Mean Age (y, SD) | Gender |
| --- | --- | --- | --- |
| Positive | 446 (14.3%) | 35.5 (10.3) | 93.0% M<br>7.0% F<br>0.0% U |
| Low Positive | 98 (3.15%) | 32.9 (11.9) | 92.9% M<br>6.1% F<br>1.0% U |
| Indeterminate | 16 (0.51%) | 33.9 (11.7) | 75.0% M<br>25.0% F<br>0.0% U |
| Negative | 2542 (81.7%) | 34.3 (18.1) | 62.7% M<br>37.1% F<br>0.2% U |
| **Total** | **3111 (100%)** | **34.4 (17.0)** | **68.2% M**<br>**31.6% F**<br>**0.2% U** |

### A Lower Repeat Threshold Improves Specificity of MPXV Detection

For samples with MPXV detected, the mean CT1 value was 20.5 (SD 4.0) for positives, 34.3 (SD 3.1) for low positives, and 37.6 (SD 3.6) for indeterminate samples. The average CT2 for low positives was 33.9 (SD 3.3). Per our algorithm, repeat testing was not performed for positive samples and CT2 values for indeterminate specimens were set at 50 (i.e. no valid curve). There was good correlation between the CT1 and CT2 of low positive specimens with an r- squared of 0.8 and standard error of 0.03 (Figure 2). During the study period a total of 184 specimens were repeated according to our algorithm. While the majority of specimens (86.4%, 159/184) detected MPXV upon repeat testing, 13.6% (25/184) of samples failed to repeat and were subsequently categorized as indeterminate. CT1 values for indeterminate specimens ranged from 29.5 to 44.2. (Figure 3). While 80% (20/25) of these samples were at or above the CDC’s recommended repeat threshold of 34, there were five specimens (CT1 values 29.5, 30.8, 31.1, 32.4, 33.3) below this cutoff. Compared to this study’s repeat algorithm, the CDC’s algorithm had a sensitivity of 100% (95% CI: 97.7% – 100%) and a specificity of 80% (95% CI: 59.3% – 93.17%) for MPXV detection in specimens with CT1 values ≥29.

**Figure 2.**
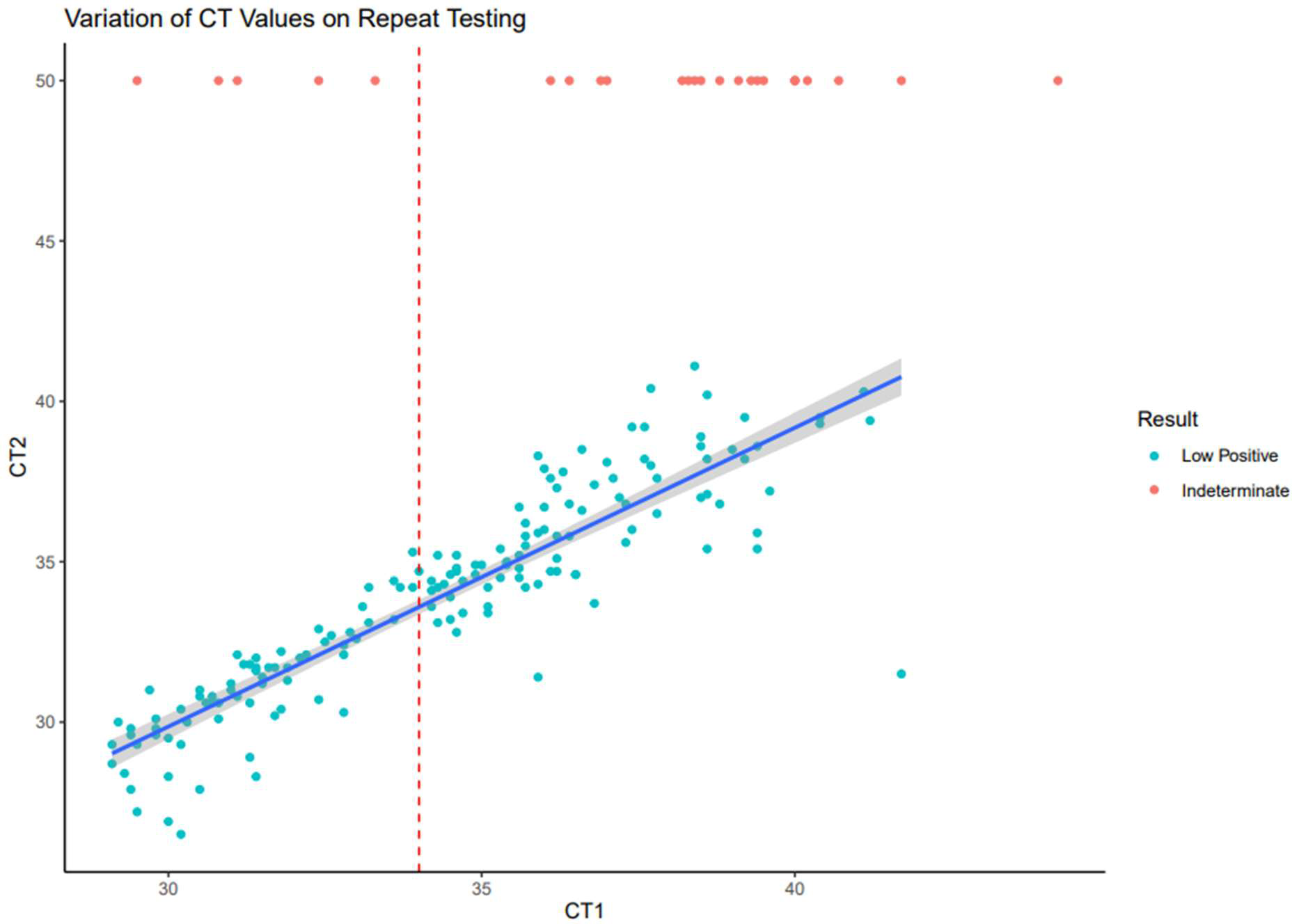
Variation of CT Values on Repeat Testing Comparison of CT values for initial (CT1) vs. repeated (CT2) tests during the study period, including both low positive results (CT1 ≥29 to <50; CT2 <50) and indeterminate results (CT1 ≥29 to <50; CT2 = 50). The CDC’s recommended repeat threshold (CT ≥34) is represented by red dotted line. Linear best fit line and 95% confidence interval for low positive results are represented by the blue line and gray shading. R-squared value for low-positive only results was 0.8. The r-squared value decreased to 0.43 when indeterminate results were included.

**Figure 3.**
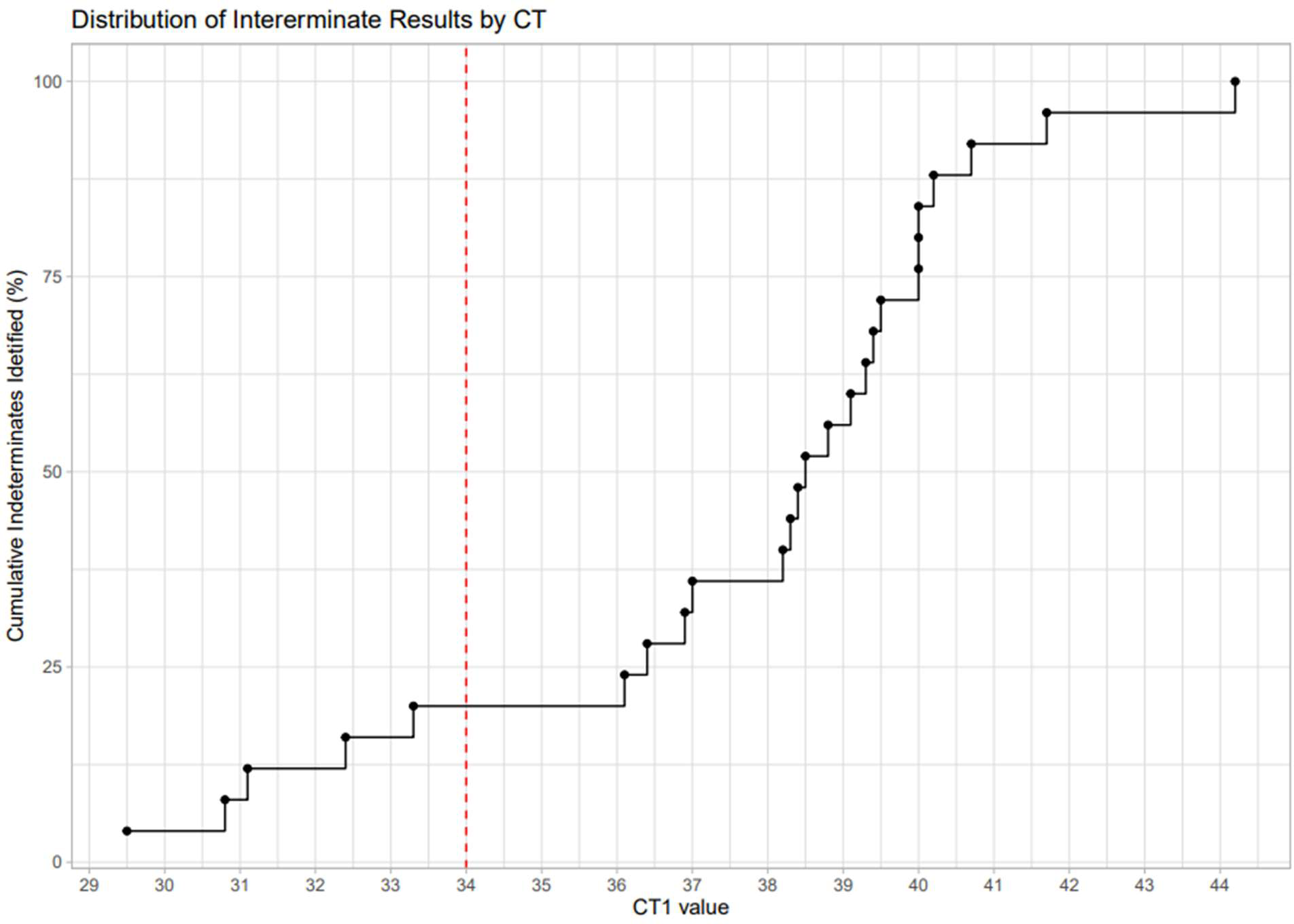
Distribution of Indeterminate Results by CT1 Cumulative percentage of indeterminate results in our study distributed by CT1 value. Each point represents one indeterminate result. Red dotted line represents the CDC’s recommended repeat testing cutoff (CT ≥34).

### A Lower Repeat Threshold Minimally Impacts Test Volume and Turn-around Times

As repeat testing could lead to unsustainable testing volume or prolonged turn-around times (TAT), we examined changes in testing volume and TAT associated with repeat thresholds of 29 or 34 as compared to a no-repeat baseline. Sub-analyses were performed assuming a mpox prevalence of 3%, 10%, 18% (the average overall detection rate during our study period), and 30%. The increase in testing volume ranged from 0.6% to 6.0% and 0.5% to 4.8% for a repeat threshold of 29 or 34, respectively (Figure 4A). During the study period, using a lower repeat threshold of 29 led to an additional 1.4 tests per day as compared to the CDC’s recommendation.

**Figure 4.**
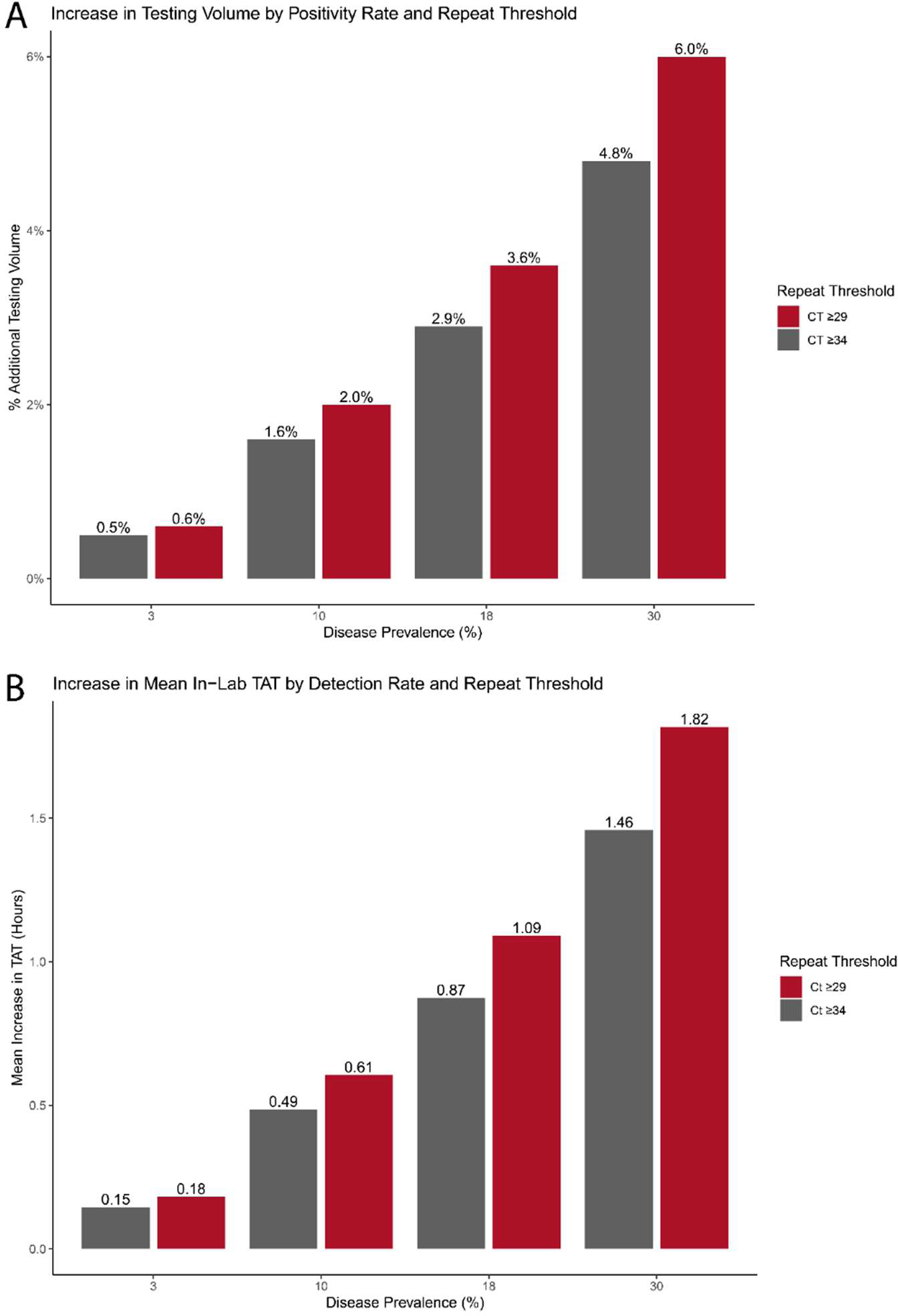
Impact of Repeat Thresholds on Overall Test Volume and In-lab TAT. A) Comparison of repeat thresholds on overall test volume based on different levels of mpox prevalence in the test population. Y-axis represent relative increase in volume versus a no-repeat approach. B) Comparison of repeat thresholds on mean in-lab TAT for specimens with MPXV detected based on prevalence. Y-axis represents the mean increase of in-lab TAT in hours versus a no-repeat approach.

TAT was assessed based on the time from specimen receipt to final result (i.e. in-lab TAT) due to variations in specimen shipping practices. Overall, the mean in-lab TAT for positive specimens increased by 0.15 to 1.82 hours depending on the prevalence of mpox and repeat threshold employed. When comparing TAT between repeat thresholds, using a CT of 29 versus 34 for repeat testing prolonged the mean in-lab TAT of positive specimens by less than 30 minutes (Figure 4B). On a per specimen basis, the average in-lab TAT was 33.1 hours (SD 16.2) for all samples, 31.9h (SD 14.2) for positives, 31.7h (SD 14.7) for negatives, 61.8h (SD 21.4) for low positives and 63.6h (SD 25.7) for indeterminate samples. Repeating a specimen added an average of 31.7h (SD 29.1) to the in-lab TAT. However, time to receipt remained the largest contributor to overall TAT at 72.8h (SD 43.0) (Figure 5).

**Figure 5.**
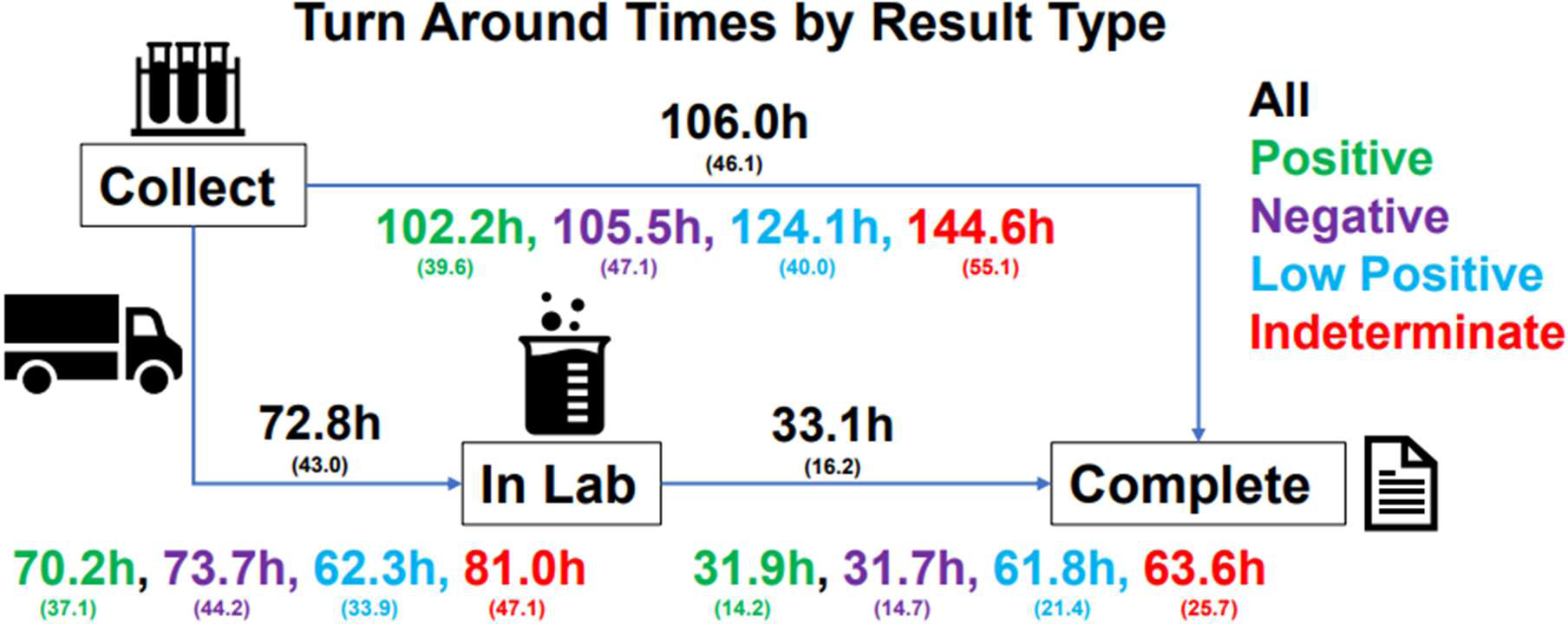
Turn Around Time by Final Result Breakdown of turn-around time by final assigned result. All times are in hours. Total time between collection and completion shown on top arrow. Values are provided as mean with standard deviation in parenthesis.

## Discussion

Here we assess over 4,000 MPXV PCR results from a large reference laboratory to examine the impact of repeating late CT specimens on patient results. We found that a reflex CT of 29 as compared to 34 potentially reduces the number of false positive results reported. We also demonstrate that regardless of the repeat threshold selected, there is minimal impact to testing volume and TAT.

False positive results may be due to a number of pre-analytic factors including mislabeling, sampling error, or contamination of collection devices and reagents^10,11^. There are also a variety of analytic errors that can occur such as pipetting error and specimen cross- contamination^12^. From the perspective of a clinical microbiology laboratory, closed tube systems and a reduction in the number of technicians handling the specimen can lower contamination rates; however, no system is perfect^13^. Specific challenges to preventing cross-contamination during the mpox epidemic included the high prevalence of positive samples (18% during the study period) and the relatively high level of MPXV DNA within positive specimens (mean CT of 20.5 for positive specimens). These problems are not unique to MPXV and may be seen in other viruses with extremely high viral loads, such as BK virus. False positive results have previously been shown with PCR assays for BK virus, including on the instrument used in this study^14^

A central question to this study is whether the specimens we categorized as indeterminate (i.e. those which failed to repeat) represent a false positive result or are an artifact of stochastic effect near the assay’s limit of detection (LOD). When validating our assay, we established the 97.5% (39/40 repeats) LOD at 80 copies per reaction which corresponded to an average CT of 39.5 (SD 1.03, 95% CI 37.8 – 41.6). Thus, we are reasonably confident that specimens at or below this CT value should consistently repeat as detected. In this study, 72% (18/25) of indeterminate specimens had a CT value less than our LOD of 39.5. Therefore, we believe the majority of indeterminate results, and in particular those identified only by the lower cutoff, represent false positive results.

Despite the CDC’s recommendation, there has been little uniformity in the adoption of MPXV repeat cutoffs. Of the 6 MPXV assays granted emergency use authorization, only one outlines a repeat strategy^15^. For this specific assay, the repeat threshold is set between a CT of 38 and 40 with the assay’s LOD determined to be at a CT of 36.0 (100 copies/mL). Selecting a repeat threshold above an assay’s LOD provides limited clinical utility as specimens which fail to repeat have a higher likelihood of representing the stochastic effect of low DNA concentrations. We selected a repeat threshold 10 cycles lower than the established LOD to more confidently identify cases which fail to repeat as false positives.

This study is unique in that it analyzes mpox specimens collected from sites across the US and includes a relatively large cohort. During the study period, we reported approximately 13% of the total number of mpox tests registered by the CDC^16^. However, these results may not translate uniformly to other laboratories. Labs with a lower throughput who have not automated this process or use a different instrument may experience different sources of error which could impact the likelihood of false positives. Limitations of this study include its retrospective and descriptive nature, use of data from a single reference laboratory, and limited clinical data to adjudicate indeterminate specimens.

While the initial surge of mpox cases has abated, the question of whether this virus will endemically circulate in humans remains^17,18^. Currently, data from the CDC and our institution show a positivity rate of approximately 10%, arguing that laboratories will continue to face testing challenges related to specimens with late CT values. As these results may have significant social and psychological impact, reducing false positive mpox results is critical^19,20^. The CDC’s recommendations provide a suitable starting point for laboratories performing mpox testing; however, each laboratory should carefully inspect the performance of its assay to determine the optimal repeat thresholds.

## Data Availability

All data produced in the present study are available upon reasonable request to the authors

